# Repeated mass testing of staff and residents in prison outbreaks of Covid-19: an enhanced outbreak investigation in two adult prisons in England, 2021

**DOI:** 10.1101/2022.06.13.22276319

**Authors:** Maciej Czachorowski, Matthew Bashton, Eamonn O’Moore, Nuala McGrath, Darren Smith, Kerry Gutridge, Julie Parkes, Emma Plugge

## Abstract

**Background:** The management of Covid-19 outbreaks presented particular challenges in the prison setting. In this study we describe the results from the implementation of a serial mass testing approach in two adult prisons in northern England. The overall aim was to examine the epidemiology of Covid-19 outbreaks in prisons and help inform public health policy and practice during the pandemic.

**Methods:** Repeat mass testing was offered to all eligible staff and residents in a women’s (n_residents_=239; n_staff_=246) and a men’s (n_residents_=703; n_staff_=340) prison in February and March 2021 at days 0, 7 and 28 after Covid-19 outbreaks were declared. Positive swab samples were sent for viral whole genome sequencing by COG-UK.

**Findings:** Participation in at least one testing round ranged from a low of 67% of staff in the men’s prison to a high of 98% of residents in the women’s prison. The largest outbreak, in the men’s prison (261 cases in residents and 37 cases in staff), continued to see new cases identified at the last testing round on day 28. Test positivity in residents of both prisons was significantly lower (p<0.05) at day 28 than on preceding test days, but no significant difference was observed for staff (p>0.05). Epidemiological data in conjunction with sequencing information provided evidence for multiple introductions of the SARS-CoV-2 virus from the local community into the prisons, with transmission identified both within wings and between wings among residents and staff. Two distinct SARS-CoV-2 lineages were identified in the women’s and men’s prisons, B.1.177 and B.1.17, respectively.

**Conclusions:** During a Covid-19 outbreak, timely implementation of a whole prison testing regime can serve to inform a targeted approach to infection prevention and control by identifying the true extent of disease transmission in all (including asymptomatic) individuals. Staff, in particular, should be tested regularly and testing uptake should be as high as possible to minimise the risk of infection incursion. Ensuring high testing uptake across all testing rounds remains a challenge.

## INTRODUCTION

The management of the COVID-19 pandemic presents particular challenges in the prison setting both because of the unique nature of the setting, which is often crowded and poorly ventilated, and also because of the prison residents who are likely to be more susceptible to severe disease due to underlying morbidities [1]. These factors can contribute to a higher risk of large and prolonged, or even recurrent COVID-19 outbreaks in prisons which are both clinically and operationally impactful if appropriate infection prevention and control (IPC) measures are not implemented early [2]. During the second wave of the pandemic, prison residents in England had a disproportionately higher age standardised mortality compared with peers in the community despite various infection prevention and control measures having been introduced, including reception testing and isolation, social distancing and compartmentalisation of populations based on infection status or likely risk from infection [3].

Critical to the timely implementation of heightened IPC measures is the need for accurate information about the number of new cases and their distribution, as identified through a dedicated outbreak testing approach. Such information provides information about the force of infection (both test positive and test negative people and where they normally reside or work in the prison) and helps to inform appropriate infection control strategy and also supports population management decisions. Repeat testing can help to understand the impact of mitigation measures, transmission networks (involving prison residents and/or staff), or loss of control of the outbreak [4].

An outbreak control team (OCT, organised by the UK Health Security Agency) will convene multidisciplinary meetings to guide the management of an outbreak in a prison in England. The OCT reviews the epidemiological evidence (including wing-specific attack rates) and will recommend further IPC measures such as restriction of movement into and out of the prison; regime restrictions to reduce social mixing; visiting restrictions to protect vulnerable visitors and reduce transmission to other prison residents; restricting staff cross-deployment across prison wings wherever possible; enhancements to recommended personal protective equipment (PPE) in affected areas; isolation of case in single-cell accommodation; prompt isolation/quarantine of possible/confirmed residents and their contacts; and the serving of meals and medicines at the door of confirmed cases to minimise movement through the prison. In addition, OCTs responding to COVID-19 outbreaks in prisons in England are encouraged to undertake a risk assessment to help decide whether to activate whole or targeted (i.e. specific wing, landing, etc.) testing (either polymerase chain reaction [PCR] and/or by lateral flow testing [LFT]) of all staff and residents working and living in a prison with a declared outbreak. At the time of the study, testing was recommended at three timepoints: at days 0 (the first day mass testing is available), between days 5 and 7, and at 28 days after the last case has been identified (i.e. after two incubation periods of SARS-CoV-2) to confirm the outbreak is over (termed recovery testing) [5].

Recommendations for mass testing during prison outbreaks were supported by intelligence that operational pressures often complicate efforts to effectively cohort staff to specific prison units and staff participation in routine asymptomatic screening programmes (weekly PCR and twice-weekly LFT at the time of the study) had fallen below recommended levels in the English prison estate [2]. While prison outbreaks often present with initial cases concentrated in specific prison wings, prisons where mass testing has been undertaken have identified COVID-19 cases in other parts of the prison suggesting the presence of transmission networks across sites even with strict IPC measures in place [6]. This informs the advice by UKHSA for whole prison testing at the point of detection of an outbreak to identify transmission networks which often involve asymptomatic or pauci-symptomatic cases.

In this study we describe the results from the implementation of a serial mass testing approach, similar to that recommended in national guidance, in two adult prisons in England that each had a COVID-19 outbreak declared in early 2021. To better ascertain disease transmission through the prison and provide information on the possible source(s) of infection, whole genome sequencing (WGS) of samples was also undertaken. The overall aim of the study was to examine the epidemiology of SARS-CoV-2 outbreaks in prisons in England and help inform public health policy and practice during the pandemic.

## METHODS

The COVID in Prisons Study (CiPS) has been described elsewhere [7]. Round 3 of the study, an enhanced outbreak investigation in two closed prisons with declared COVID-19 outbreaks in February 2021, is detailed in this report. Briefly, outbreaks of COVID-19 were declared by local UKHSA Health Protection Teams (HPTs) in a number of the 28 prisons participating in the CiPS study in early 2021. The first two prisons in which an outbreak was declared, and mass testing was operationally feasible, were approached to participate. Case-level symptomatology data was not available for staff or residents as part of the mass-testing programme.

Participating establishments included a women’s prison and a men’s prison both located in the north of England. The women’s prison has a capacity of over 300, and the men’s prison of over 700 people. All voluntarily consenting residents aged 18 years or older were offered throat and nasopharyngeal swabs at approximately 0, 7 and 28 days after each outbreak had been declared by the local HPT. Directly and non-directly employed prison staff were also asked to consent to share data for the study collected as part of ongoing twice weekly ‘business as usual’ staff testing undertaken during the study period.

Swab samples from participating staff and residents were sent to regional labs for polymerase chain reaction (PCR) analysis on which COVID-19 diagnosis was based (positive/negative); test results were available within 72 hours. Trained private contractors were used to test residents (researchers were not allowed into prisons during the pandemic). At first contact, each resident provided written consent and at subsequent testing points, verbal consent was ascertained. The swab was self-administered by the residents with supervision from the trained contractors to ensure that it was performed correctly and to safely collect the sample. Participants who had previously tested positive for COVID-19 in the 90 days prior to testing did not receive a PCR test, as per national guidance [5]. Staff who were shielding (because clinically extremely vulnerable) or on long-term absences for the duration of the pandemic were excluded from testing.

Positive samples were couriered securely to a Coronavirus Disease 2019 (COVID-19) Genomics UK Consortium (COG-UK; a national, multi-centre consortium for the sequencing and analysis of SARS-CoV-2 genomes for Public Health) study laboratory for genomic sequencing of the virus [8]. The background data from the UK was generated by the COG consortium (https://www.cogconsortium.uk/). The phylogenetic analysis was done using CIVET version 2.0 https://github.com/artic-network/civet.

Demographic information was collected from prison-National Offender Management Information System (p-NOMIS) records [9] for participating residents and from electronic staff records which included information about COVID-19 symptom presentation (at time of testing). Staff and resident population numbers for each participating establishment, used to derive participation proportions at the time of testing, were obtained from the Ministry of Justice Offender and Workforce population statistics [10, 11].

Frequencies and percentages were used to describe categorical variables and medians and ranges were used to describe participant age variables. The Clopper-Pearson exact method was used to calculate 95% confidence intervals for testing proportions [12]. Relevant statistical tests at a significance level of 0.05 were used to evaluate associations between variables. As most participants only participated in a single testing round (Table 1), independence between testing rounds was assumed for the purposes of statistical comparisons of test positivity. All analyses were performed using STATA statistical software, version 15.1.

**Table 1:** Study participation by prison, participant cohort and number of study rounds

| Prison | Cohort | Eligible prison pop.* | Study participants† | Overall participation† | Number of testing rounds completed by study participants |  |  |  |  |  |
| --- | --- | --- | --- | --- | --- | --- | --- | --- | --- | --- |
|  |  |  |  |  | 1 round only |  | 2 rounds |  | all 3 rounds |  |
|  |  |  |  |  | n | % | n | % | n | % |
| Men's | Residents | 703 | 499 | 71.0 | 292 | 58.5 | 125 | 25.1 | 82 | 16.4 |
|  | Staff | 340 | 227 | 66.8 | 182 | 80.2 | 38 | 16.7 | 7 | 3.1 |
| Women's | Residents | 239 | 234 | 97.9 | 83 | 35.5 | 78 | 33.3 | 73 | 31.2 |
|  | Staff | 246 | 206 | 83.7 | 122 | 59.2 | 61 | 29.6 | 23 | 11.2 |

\*excludes individuals who were swabbed, and later tested positive, in pre-study period
†participated (i.e. were tested) in at least one testing round e.g. 'Day 0, 7 or 28'

## RESULTS

### 1. Epidemiological data

Overall study participation was good in both residents and staff with more than 66% of the eligible population participating in the men’s prison and more than 83% in the women’s prison (Table 1). However, participation in more than one testing round was low, with the majority of participants in both prisons having only been tested once rather than at each of the ‘Day 0, 7 and 28’ testing rounds (Table 1). Participant characteristics by cohort and prison are detailed in Table S1.

### Men’s prison

The men’s prison comprised ten wings with most resident cases having been detected on a single wing (C-wing) prior to mass-testing taking place. The outbreak was declared on February 22^nd^ and mass testing took place from February 25^th^ to March 28^th^ (Figure 1A). In the ten days before mass testing, 85 cases were swabbed in residents (32.6% of all positive tests; 85/261) and 31 in staff (83.8% of all positive tests; 31/37) (Table S2). Test uptake peaked in residents at the second test round, ‘Day 7’, with 48.4% of eligible men participating (Table S2). In contrast, staff participation was highest at the last test point, ‘Day 28’, during which 56.3% of eligible staff participated (Table S2). New cases of COVID-19 were confirmed throughout the testing period in residents, but only at ‘Days 7 and 28’ in staff (Figure 1A and Tables 2 and S2). Furthermore, test positivity was significantly lower (p<0.05) in residents at the last test point, ‘Day 28’ (14.8%; 10.4%-20.1%), than at either of the two prior test points, ‘Day 0’ (27.5%; 21.8%-33.8%) and ‘Day 7’ (25.9%; 21.1%-31.2%) (Table 2).

**Table 2:** Positivity rates based on total valid tests by testing round and participant cohort in each study prison

| Prison | Cohort | 'Day 0' |  |  |  | 'Day 7' |  |  |  | 'Day 28' |  |  |  | Total valid tests* | p-val.† |
| --- | --- | --- | --- | --- | --- | --- | --- | --- | --- | --- | --- | --- | --- | --- | --- |
|  |  | Positive tests | Valid tests* | Positivity | 95% CI | Positive tests | Valid tests* | Positivity | 95% CI | Positive tests | Valid tests* | Positivity | 95% CI |  |  |
| Men's | Residents | 63 | 229 | 0.275 | 0.218 0.338 | 80 | 309 | 0.259 | 0.211 0.312 | 33 | 223 | 0.148 | 0.104 0.201 | 761 | **0.001** |
|  | Staff | 0 | 46 | 0.000 | 0.000 0.077§ | 1 | 42 | 0.024 | 0.001 0.126 | 5 | 191 | 0.026 | 0.009 0.060 | 279 | 0.684 |
| Women's | Residents | 9 | 132 | 0.068 | 0.032 0.125 | 2 | 174 | 0.011 | 0.001 0.041 | 0 | 148 | 0.000 | 0.000 0.025§ | 454 | **0.0003** |
|  | Staff | 2 | 158 | 0.013 | 0.002 0.045 | 0 | 88 | 0.000 | 0.000 0.041§ | 0 | 65 | 0.000 | 0.000 0.055§ | 311 | 0.712 |

\*excludes void results and non-returns
† Fisher's exact test for difference in proportions across testing rounds; bold values statistically significant
§ one-sided, 97.5% confidence interval

**Figure 1:**
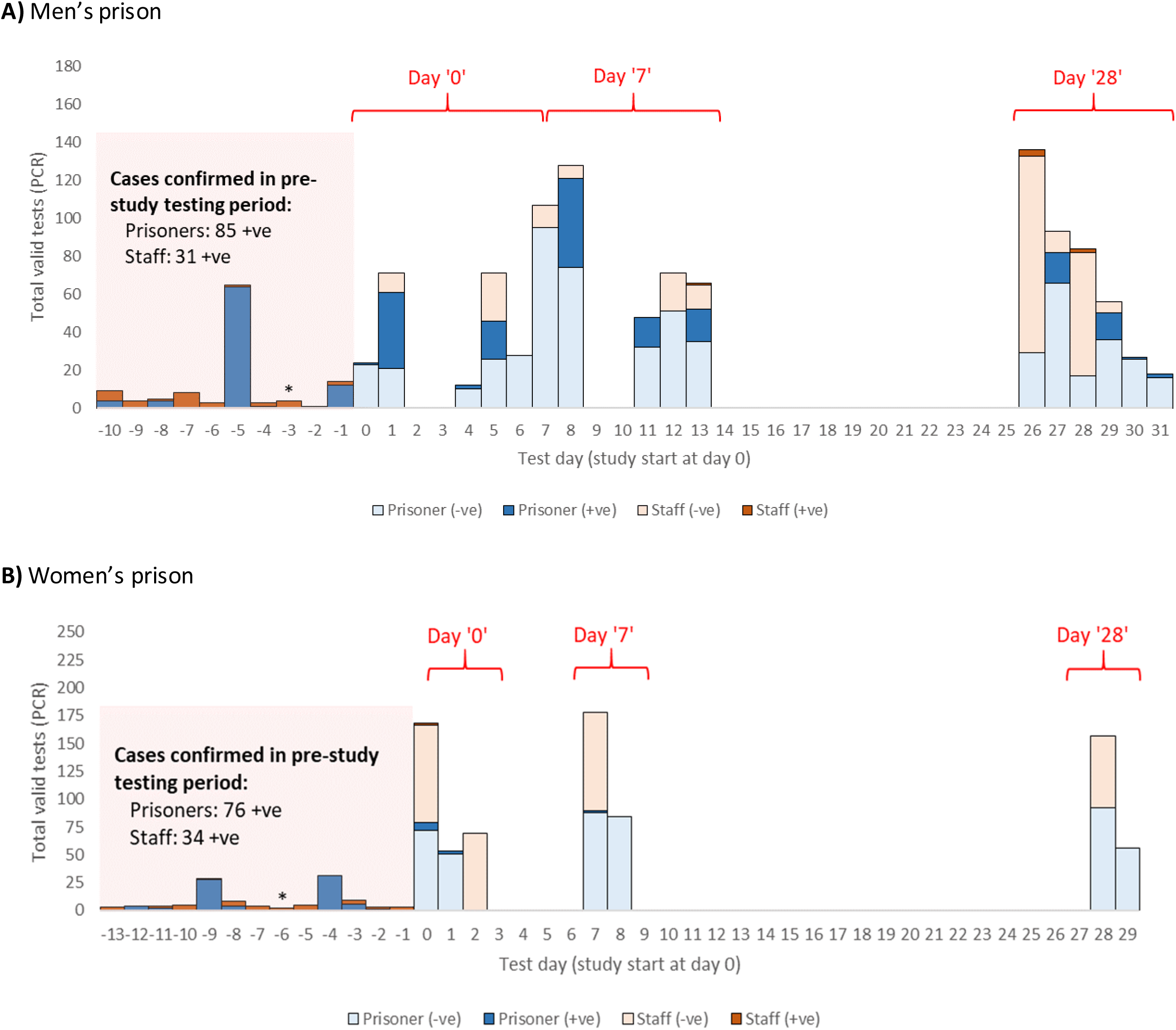
Histograms of total valid polymerase chain reaction (PCR) tests for SARS-CoV-2 in staff and residents by study prison and testing rounds (‘Days 0, 7 and 28’). In the ‘pre-study’ testing period (shaded region in histograms, below), only data on the total number of positive tests was available. Asterisk (*) represents the day on which outbreak was officially declared by outbreak control team (OCT).

Cases in residents were detected across seven prison wings (Table S3) and positivity rates varied considerably by location within the prison; data on staff deployment were not available. With the exception of D-wing, where test positivity continued to increase at each successive test round (significantly so from ‘Day 7’ to ‘Day 28’), test positivity on the affected wings was lower by the last test round, ‘Day 28’, than at preceding test rounds i.e. ‘Days 0 and/or 7’ (Table S3). No significant difference in test positivity amongst the three test rounds was observed in staff (Table 2).

### Women’s prison

The outbreak in the women’s prison was declared on February 9^th^ at which point IPC measures were introduced and three rounds of mass testing took place from February 15^th^ to March 16^th^ (Figure 1B). The majority of cases were swabbed in the 13-day period before mass testing took place: 76 in residents (86.4% of all positive tests; 76/88) and 34 in staff (94.4% of all positive tests; 34/36) (Table S1). At the time of the study, the prison had 315 residents and 280 staff (Table S2); approximately 30% of resident cases had COVID-19 symptoms when tested (data not shown). The proportion of staff participating in testing dropped following each successive test point, from a high of 64.2% at ‘Day 0’, to a low of 26.6% at ‘Day 28’ (Table S2). In contrast, test participation peaked at ‘Day 7’ in residents, with 76.1% of eligible women participating (Table S2). No new cases were identified in participating residents or staff at the last testing round i.e. at ‘Day 28’.

In residents, test positivity was markedly lower at each successive test round and dropped significantly from a high of 6.8% (95% CI: 3.2% - 12.5%) at ‘Day 0’ to 0.0% (95% CI: 0.0% - 2.5%) at ‘Day 28’ (Table 2). Workforce constraints limited the number of prison wings that could be tested at any one test round, but cases were identified across three wings (B, 3 cases in total; C, 2 cases in total; J, 6 cases in total) (Table S3). Given the low case numbers, no significant differences (p>0.05) in test positivity were observed between test rounds in residents or staff (Table S3).

Incidence rates were calculated in residents in both prisons but could not be calculated in staff as so few staff participated in more than one sweep of testing. Incidence rates, in line with test positivity, declined in subsequent test rounds (compare incidence rates from ‘Day 0 and 7 tests’ to ‘Day 7 and 28 tests’ in Table 3).

**Table 3:** COVID-19 incidence rates by follow-up period in residents of each study prison

| Prison | Follow-up period | Follow-up time (person/weeks) | New cases | Incidence (cases/person/week) | 95% CI |
| --- | --- | --- | --- | --- | --- |
| Women's | Day 0 to 7 | 104.784 | 2 | 0.019 | 0.005 - 0.076 |
|  | Day 7 to 28 | 322.466 | 0 | 0.000 |  |
|  | Day 0 to 28 | 462.842 | 2 | 0.004 | 0.001 - 0.017 |
| Men's | Day 0 to 7 | 96.384 | 10 | 0.104 | 0.056 - 0.193 |
|  | Day 7 to 28 | 330.439 | 27 | 0.082 | 0.056 - 0.119 |
|  | Day 0 to 28 | 525.342 | 37 | 0.070 | 0.051 - 0.097 |

### 2. Genomic analysis

Of the 210 positive swab samples collected from staff and residents as part of mass testing in the women’s and men’s prisons, 177 (84.3%) were sent to the COG-UK laboratory for WGS, and 121 (68.4%) of these were successfully sequenced representing 57.6% of all positive samples collected.

### Women’s prison

Of the 12 positive samples identified in residents during the mass testing phase in the women’s prison, 9 (75%) were successfully sequenced, as were both (100%) positive samples in staff. All sequenced samples collected in the prison were part of the same viral sub-lineage, B.1.177.10, derived from a lineage (B.1.177) that emerged in early summer 2020, probably in Spain, and subsequently spread to multiple locations in Europe, including the United Kingdom [13]. A phylogenetic analysis (Fig. 2) of these sequences indicates multiple phylotypes were linked to the outbreak, often with only a single nucleotide polymorphism (SNP) difference between them. Two prominent but distinct phylogenetic clusters were seen in several cases resident on B and J wings. These two phylogenetic clusters, along with two other unique viral sequences identified in residents on C wing, appear to branch from a common viral sequence identified in a resident on J wing (Fig. 2), suggestive of inter-wing viral transmission from resident(s) on J wing directly (e.g. resident to resident) or indirectly (e.g. via staff) to residents on B and C wings. The identical nature of the sequenced genomes within the B and J wing branches of the tree is also suggestive of intra-wing viral transmission among residents of these twowings following the initial acquisition event. Resident-staff transmission is also clearly evidenced by inclusion of a member of kitchen staff in the same phylogenetic cluster seen primarily on J wing. However, the presence of three collapsed community nodes in the phylogenetic tree, suggests that multiple introductions into the prison of the viral sub-lineages observed cannot be ruled out (grey squares in Figure 2). The sequence from the second staff member to test positive, did not map to the phylogenetic tree shown in Figure 2 and likely resulted from community acquired infection (not shown).

**Figure 2:**
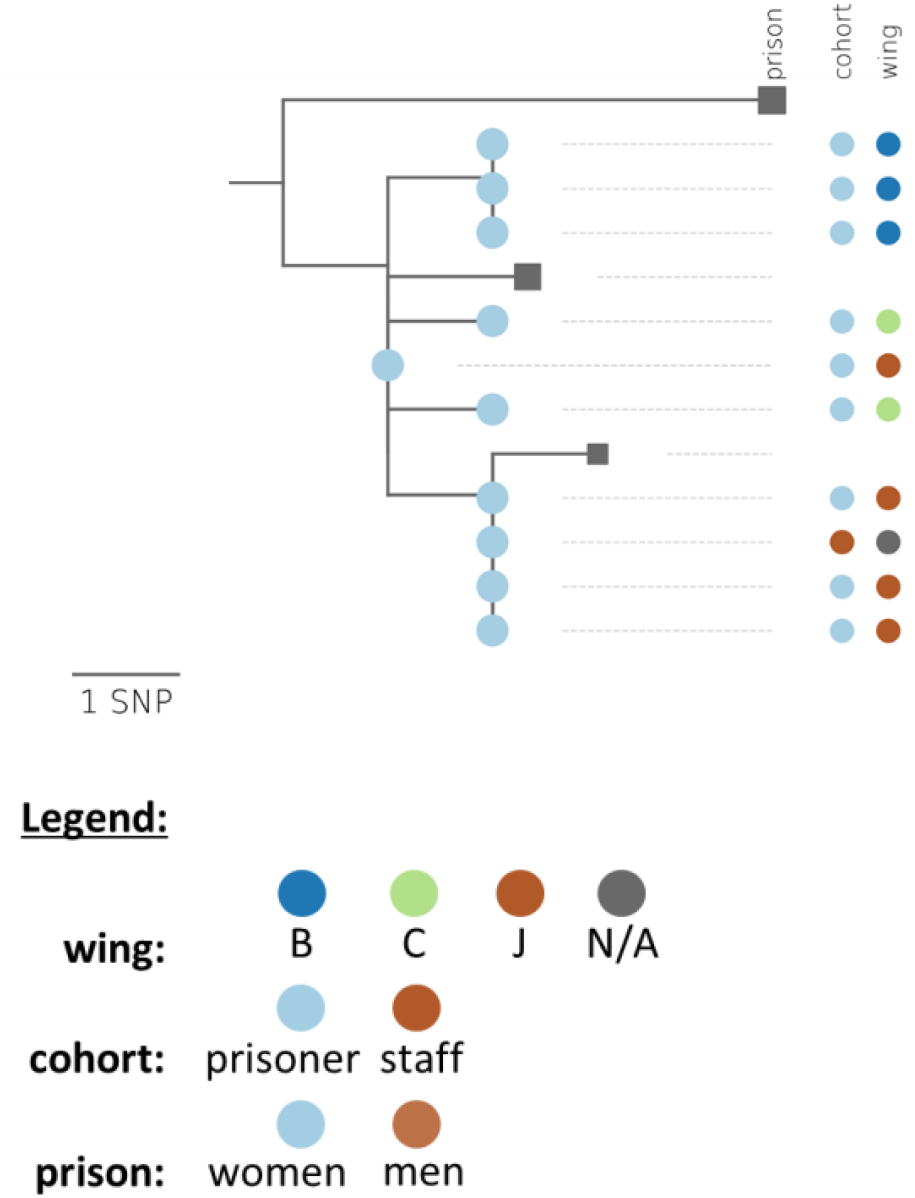
Phylogenetic tree showing genetic distance between available SARS-CoV-2 specimens from the women’s prison. Related community clusters interspersed among prison samples also shown as collapsed nodes represented by grey squares. Scale bar represents a difference of a single nucleotide polymorphism (SNP). N/A = information not available.

### Men’s prison

Of the 190 positive swab samples collected from residents as part of mass testing in the men’s prison, 157 (82.6%) were sent to the COG-UK laboratory for WGS, and 106 (67.5%) of these were successfully sequenced, representing 55.8% of all positive samples from residents. In staff, four (66.7%) of the six positive samples that were identified by mass-testing were sent for sequencing and successfully sequenced. All sequenced samples in the men’s prison were derived from a UK phylotype (UK1726_1.1.2.468.309.10) of the B.1.1.7 lineage of SARS-CoV-2, known for its high transmissibility and also a known variant of concern (VOC) [14]. Various viral sub-phylotypes are shown tobranch from this lineage in the main phylogenetic tree which maps 109 cases (105 residents and 4 staff) in the men’s prison (Figure 3). Several regions of phylogeny are shown to localise to specific prison wings, and sequencing within these clusters revealed the acquisition of extra mutations, suggesting high levels of intra-wing viral transmission e.g. phylotypes clustering on C-, D-, F- and H-wings (Figure 3). The two most prominent polytomous branches of the phylogenetic tree contain viral sequences from residents residing on 1) C and D wings (all descended from the H-wing cluster), and 2) C and G wings, suggestive of inter-wing viral transmission. Interspersed throughout the phylogenetic tree are several collapsed nodes with viral phylotypes homologous to those found in the community, suggesting that multiple importation events are likely to have contributed to this outbreak (Figure 3; see grey squares). Furthermore, sequences homologous to the central COG-UK phylogenetic tree also appear in Figure 3 as small grey circles, and likely result from directly linked cases, missing samples from the study or individuals who were also tested in the community in addition to participating in the study i.e. duplicates. One sequence from a resident on D-wing did not map to the main phylogenetic tree linked to this outbreak, suggesting community acquired infection in this individual (presumably via recent visitor contact or new admission; resident admission information was not provided for this case) and no onward transmission (not shown).

**Figure 3:**
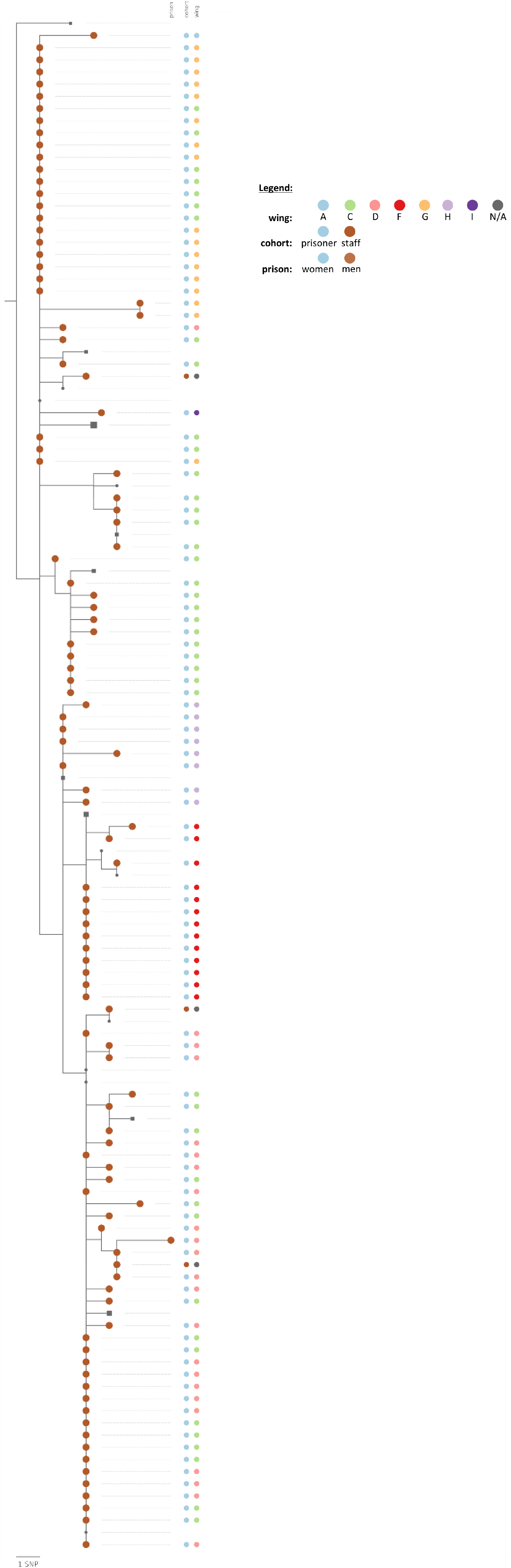
Phylogenetic tree showing genetic distance between available SARS-CoV-2 specimens from the men’s prison. Related community clusters interspersed among prison samples also shown as collapsed nodes represented by grey squares. Homologous sequences from central COG-UK phylogenetic tree shown as grey circles. Scale bar represents a difference of a single nucleotide polymorphism (SNP).

## DISCUSSION

In this study we examined the real-world implementation of sequential mass SARS-CoV-2 testing as a means of informing outbreak response in two distinct prisons in England: a women’s prison and a men’s prison. WGS data from cases was used in conjunction with epidemiological data to better understand transmission dynamics in the two adult prisons. Aside from distinct resident populations, the size, duration and extent of the outbreaks reported in each prison also substantially differed.

It was logistically challenging to rapidly mobilise testing of the large populations in each prison and mass testing at ‘Day 0’ occurred from 3-6 days after the outbreaks had been declared. Such delayed mobilisation of the testing workforce resulted in a sizeable proportion of people who tested positive for SARS-CoV-2 being identified before the study testing period had started. The advice to implement mass, or targeted, testing during a prison outbreak is national health policy in England and directed by the OCT. Arrangements to support timely outbreak testing within prison and youth custody settings was agreed between the prison service (HMPPS), the health service (NHSE/I), the Department of Health (DHSC) and the national public health agency (PHE). Testing requests are made via submission of a completed proforma to HMPPS and testing is conducted via community testing laboratories using the prison estate’s supply of test kits. This process was implemented in August 2021 across the English prison estate and aims to reduce the implementation time of mass or targeted testing in prisons to no more than 48 hours from the initial request.

Although case-level data was not collected, prison health care teams reported that a large number of cases identified in the study were asymptomatic (about two-thirds of cases in the women’s prison and nearly all those in the men’s prison). OCTs are able to consider evidence of the specific prison incident to guide testing but most prison outbreaks last for many weeks and contributing to this is the delay in getting clear understanding of the force of infection and interrupting transmission networks effectively. This highlights the need to test the whole prison to understand the true extent of an outbreak. Relying on identification of infection in other parts of a prison purely through the identification of symptomatic cases risks losing control of an outbreak as the majority of cases in residents and staff to date have been asymptomatic [6]. This is particularly pertinent in a prison setting where residents may often have incentives not to report symptoms given the restrictions to freedoms that isolation inherently entails [1].

Overall, test positivity in residents at both study prisons was significantly lower at ‘Day 28’ than at the ‘Day 0’ testing point, however cases were still detected at the later point. Furthermore, despite the implementation of IPC measures, unaccounted for inter-wing transmission among (presumably asymptomatic) residents can ‘seed’ isolated outbreaks on previously unaffected prison wings as overall positivity begins to subside. This was observed in the men’s prison which recorded a significant increase in SARS-CoV-2 positivity from ‘Day 7’ to ‘Day 28’ despite no cases being identified at the first test point, ‘Day 0’. These findings support the importance of prison wide ‘recovery testing’ from one to two incubation periods after the last case(s) are reported in a prison outbreak. Recovery testing also helps ensure that asymptomatic transmission does not continue to drive infection just as IPC measures are set to be relaxed following closure of the outbreak. At the time of this study, recovery testing was recommended in prisons two incubations periods (i.e. 28 days) after the last case, but in February 2022 this was reduced to one incubation period (i.e. 14 days) on the balance of operational pressures across the estate and new information about the biology of the omicron variant that was most prevalent in England at the time.

The genomic analysis revealed that multiple rather than single introductions of infection into the prison occur during an outbreak, most likely from the local community. It was not possible to determine with certainty whether these infections were brought in by staff or newly arriving residents living locally; this is unlikely in the case of the women’s prison which holds women from all over the country. The findings highlight the importance of regular staff testing to identify infected staff as they are the most likely vector unknowingly bringing infections into prisons during an outbreak, reflect the level of infection in their local community, and have a high degree of social mixing with other staff and with residents. Staff uptake of testing should be as high as possible to minimise the risk of infection incursion into prisons - UKHSA have recommended that at least 75% adherence to the prescribedtesting protocol is consistently required across staff groups. This is particularly important given that staff positivity did not significantly differ at any of the three test points in the men’s prison, despite a fall in positivity by the last test point in residents. Test uptake rates were higher for residents with up to 74% of residents agreeing to be tested, and consistently lower in staff with a low of 12% of staff participating in one testing round.

When test uptake rates are low, infected staff will not be identified and pose a risk to colleagues and those detained. The very low rates of testing uptake amongst staff, even in outbreaks, is of considerable concern. Work in care homes which are similar but not entirely comparable settings, demonstrated the clear role of staff in transmitting infection to residents within and between care homes [15]. Care home staff worked across different care homes, and it is likely that infection was spread between care homes in this way. This is not common practice for prison officers. Non-adherence with asymptomatic testing protocols by staff in prisons may be driven by several factors including perceived risks of infection by staff or of transmission with serious consequences for them or the people in their care, lack of leadership in requiring testing protocols to be adhered to strictly, or financial incentives not to test which could result in financial penalties like not being able to work overtime. To improve testing adherence amongst prison staff, a policy change was made in the English prison estate on December 23, 2021 mandating COVID-19 testing for all prison staff.

Evidence of cases in staff working across more than one wing and/or evidence of infection in residents in more than one wing should prompt testing at pace and scale. OCTs should consider recommending that the prison stops cross-deployment of staff within the prison as untested staff who even though infected are likely to be asymptomatic can act as vectors of transmission within (as well as into) the prison. Residents being received into prison from the community also pose an infection risk and it is important to maintain strict IPC measures on reverse cohorting units where they are quarantined prior to joining the general prison population.

### Strengths and limitations

To the best of our knowledge, this is the first national study to examine sequential mass testing for SARS-CoV-2 in prisons with declared outbreaks. It provides valuable intelligence to inform practice, public health and policy making. However, there were a number of limitations with the study. As already noted, testing uptake rates were low, particularly among staff, and it is likely that the positivity rates are an underestimate of the true prevalence. Furthermore only 57.6% of all positive samples were successfully sequenced and linked and therefore the genomic analysis was able to provide a partial picture only. However, the available WSG findings provided unique and valuable information showing introduction of infection from the community with intra- and inter- wing transmission despite the fact that OCT meetings minutes recorded strict IPC measures.

## CONCLUSIONS

In an outbreak situation, mass testing of the whole prison, not just the affected wing(s), is important to identify the true extent of the outbreak which is unlikely to be confined to the wing(s) already identified as having test positive and/or symptomatic prison residents. Relying on patient reported symptoms alone to identify infected and therefore infectious individuals is not sufficient especially in a prison setting where there may be cultural and behavioural barriers to self-reporting due to consequences for the individual and their cell-mates or others, such as isolation and lack of access to the prison regime, including visiting. Regular pre-shift staff testing is important to identify infected staff as they are the most likely vector bringing infections into prisons. Staff uptake of testing should be as high as possible to minimise the risk of infection incursion into prisons.

## Data Availability

All data produced in the present study are available upon reasonable request to the authors.

## SUPPLEMENTARY TABLES

**Table S1:**
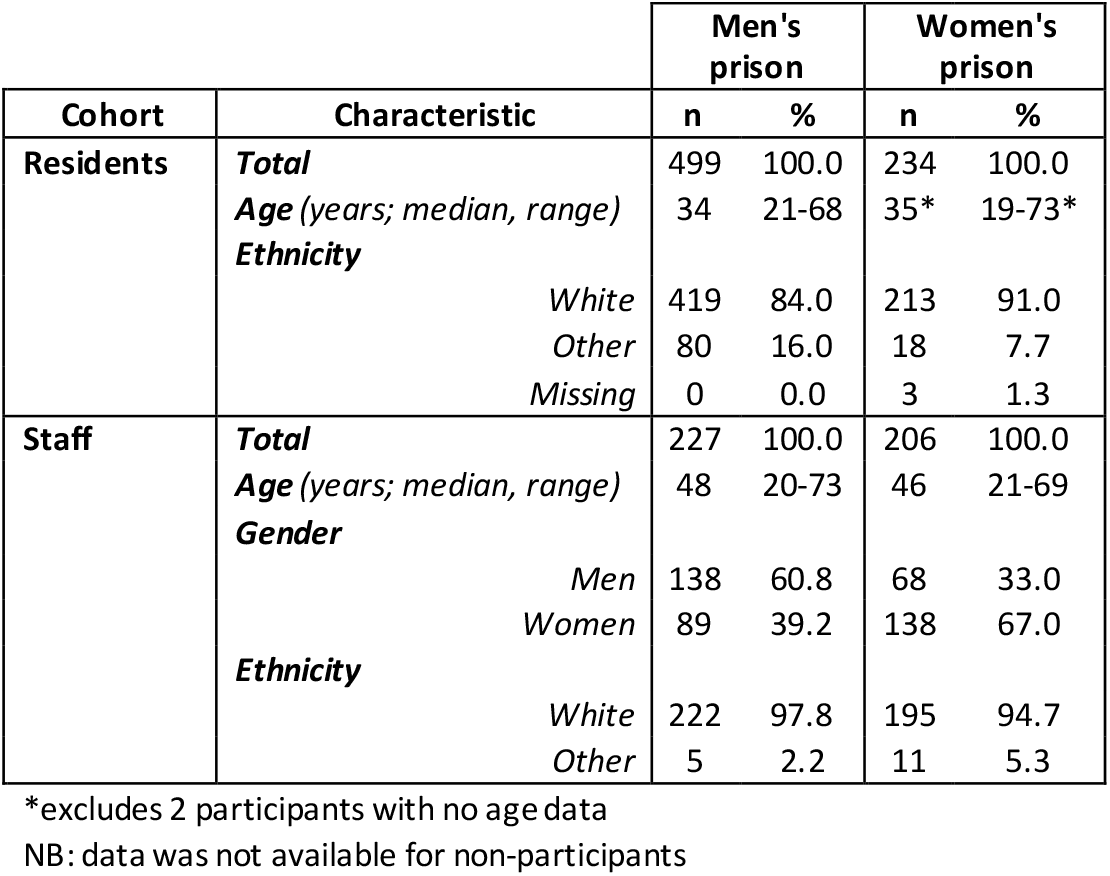
Participant characteristics in both study prisons

**Table S2:** Participation rates in each study prison by testing round and participant cohort

| Prison | Cohort | Total prison pop. | Positive cases (pre-study period) | 'Day 0' |  |  | 'Day 7' |  |  | 'Day 28' |  |  | Total tests |
| --- | --- | --- | --- | --- | --- | --- | --- | --- | --- | --- | --- | --- | --- |
|  |  |  |  | Eligible* n | Tested n | Tested % | Eligible* n | Tested n | Tested % | Eligible* n | Tested n | Tested % |  |
| Men's | **Residents** | 788 | 85 | 703 | 250 | 35.6 | 640 | 310 | 48.4 | 560 | 228 | 40.7 | 788 |
|  | **Staff** | 371 | 31 | 340 | 46 | 13.5 | 340 | 42 | 12.4 | 339 | 191 | 56.3 | 279 |
| Women's | **Residents** | 315 | 76 | 239 | 135 | 56.5 | 230 | 175 | 76.1 | 228 | 148 | 64.9 | 458 |
|  | **Staff** | 280 | 34 | 246 | 158 | 64.2 | 244 | 90 | 36.9 | 244 | 65 | 26.6 | 313 |

\*participants who tested positive within previous 90 days were not eligible for testing and excluded from eligible population

**Table S3:**
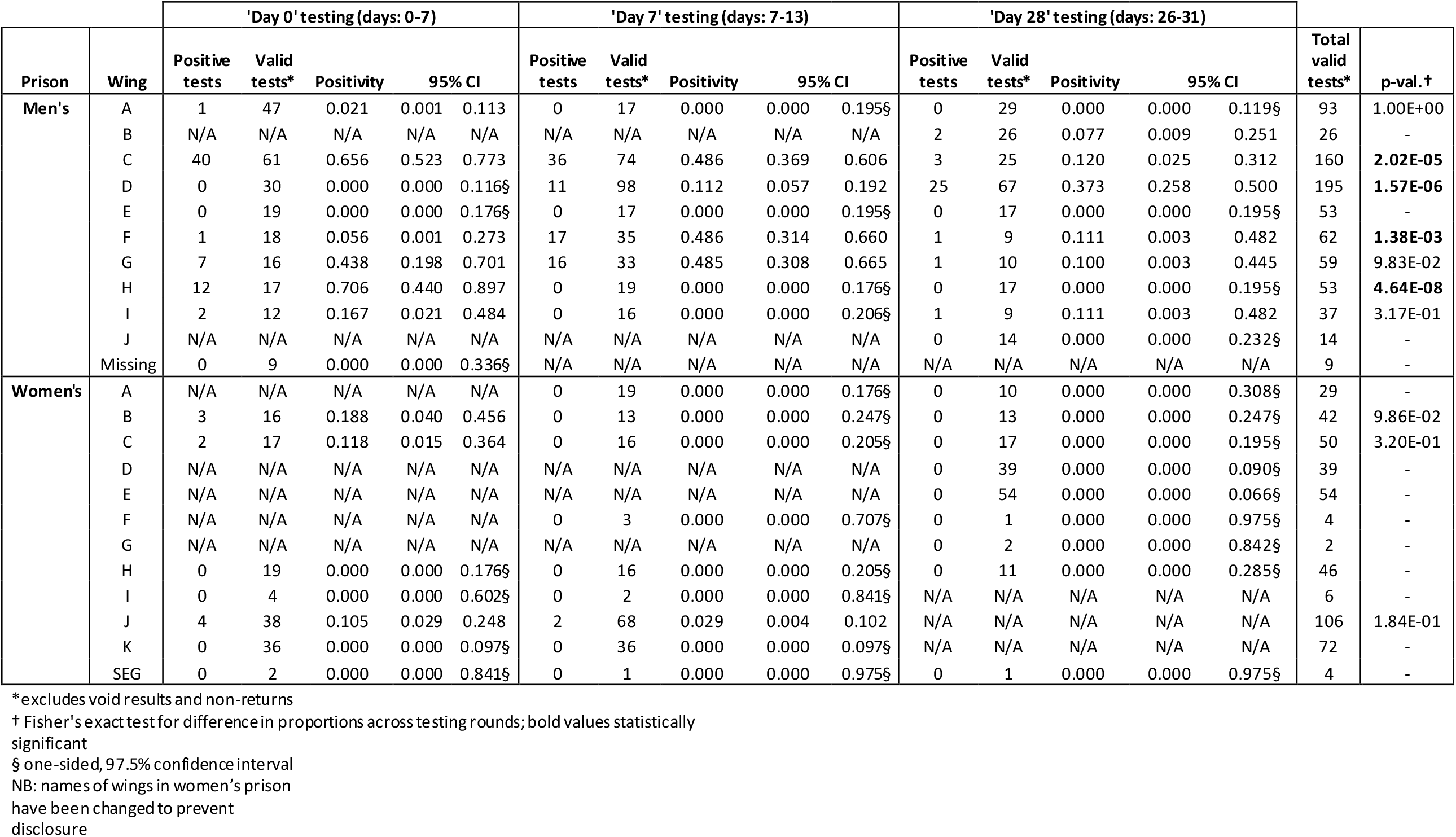
SARS-CoV-2 positivity by testing round (0, 7 and 28) and prison wing in both study prisons

